# CRISPR-based multiplex detection of human papillomaviruses for one-pot point-of-care diagnostics

**DOI:** 10.1101/2023.12.04.23299381

**Authors:** Ahmed Ghouneimy, Zahir Ali, Rashid Aman, Wenjun Jiang, Mustapha Aouida, Magdy Mahfouz

**Affiliations:** Laboratory for Genome Engineering and Synthetic Biology, Division of Biological Sciences, 4700 King Abdullah University of Science and Technology, Thuwal 23955-6900, Saudi Arabia; Division of Biological and Biomedical Sciences, College of Health and Life Sciences, Hamad Bin Khalifa University, Education City, Qatar Foundation, Doha, Qatar, P.O. Box: 34110)

**Keywords:** CRISPR-Cas, Human Papillomavirus, Sexually transmitted infections, Molecular diagnostics, Multiplex detection, Point-of-Care detection, Cervical cancer

## Abstract

The World Health Organization’s global initiative toward eliminating high-risk Human Papillomavirus (hrHPV)-related cancers recommends DNA testing over visual inspection in all settings for primary cancer screening and HPV eradication by 2100. However, multiple hrHPV types cause different types of cancers, and there is a pressing need for a, easy-to-use, multiplex point-of-care diagnostic platform for detecting different hrHPV types. Recently, CRISPR-Cas systems have been repurposed for point-of-care detection. Here, we established a <u>CRISPR</u>-Cas multiplexed <u>d</u>iagnostic assay (CRISPRD) to detect cervical cancer-causing hrHPVs in one reaction (one-pot assay). We harnessed the compatibility of thermostable AapCas12b, TccCas13a, and HheCas13a nucleases with isothermal amplification and successfully detected HPV16 and HPV18, along with an internal control in a single-pot assay with a limit of detection of 10 copies and 100% specificity. This platform offers a rapid and practical solution for multiplex detection of hrHPVs, facilitating large-scale hrHPV point-of-care screening, guiding effective treatment and policy development, and supporting global health initiatives. Furthermore, the CRISPRD platform’s programmability enables it to be adapted for simultaneously detecting hrHPVs-causing cancer, strengthening the scope of early diagnosis of HPV in the fight against infectious disease.

## Introduction

Globally, human papillomavirus (HPV) infections contribute to approximately 4.5% of all malignancies, accounting for 8.6% of cancer cases in women (the third most prevalent cause with a high mortality) and 0.8% of cases in males^1–3^. In 2020, nearly 600,000 women were diagnosed with cervical cancer worldwide, leading to 300,000 associated deaths^4^. Based on the severity of this preventable cancer, the World Health Organization (WHO) has issued a global 90-70-90 call-to-action, setting clear targets to reduce cervical cancer by 2030 and irradicate HPV by 2100 through increasing HPV vaccination to 90% of the world’s population, implementing twice-lifetime screening in 70% of the female population, and treating 90% of cervical cancer patients^5^. In developed countries, government-funded cancer screening and HPV vaccination programs have reduced the incidence and mortality of cervical cancer. However, in less-developed countries, cervical cancer remains a leading cause of cancer-related deaths^6^.

HPV infection is transmitted through sexual contact, and it is estimated that 80% of the sexually active population will experience at least one infection of HPV in their lifespan. Persistent infection by high-risk HPV (hrHPV) strain(s) and expression of HPV oncogenes leads to cervical intraepithelial neoplasia (CIN). Cervical disease is classified into three stages: CIN1 represents the transient infection by HPV and has a high probability (90%) of HPV regression and clearance. CIN2 is the next stage, characterized by persistent replication but unproductive infection in the basal layer. CIN3 is the high-grade precancerous stage, and if not regressed, epithelial-layer malignancy leads to cervical cancer^7–9^. The progressive nature of HPV infection and CIN provides an HPV-related cancer-prevention window as long as HPV is detected early and cervical disease is intercepted at the initial stages (CIN1 or CIN2).

The replicative genome of HPV consists of a double-stranded circular DNA of approximately 7900 base pairs. The genome contains eight overlapping open reading frames: six early (E) genes, two late (L) genes. The E1 and E2 genes encode proteins involved in regulating HPV replication and transcription of early proteins, and the E6 and E7 genes encode oncoproteins^10^. The L1 and L2 genes encode the major and minor capsid proteins, respectively. Persistent hrHPV infection leads to the constitutive expression of E6 and E7, which are responsible for host cell cycle regulation via controlling p53, pRB and DNA repair inhibition, ultimately establishing cancer hallmarks and tumorigenesis at the upper cervix cell layer^11^. The two most oncogenic hrHPV genotypes are HPV16 and HPV18, correlated with 95% of HPV-positive cervical cancer cases^12^. Early stages of hrHPV-associated cervical cancer are asymptomatic, making HPV testing with subtyping for HPV16 and HPV18 E6 and E7 targets a valuable primary screening tool with higher throughput, sensitivity, and reproducibility than standard cytology tests^13^.

Early detection of hrHPVs and associated malignancies is crucial for guiding effective preventative strategies against hrHPV-related cancers. Large-scale self-testing programs can substantially limit the spread of the virus, reduce infection rates, and facilitate early prognosis of cancer development. However, current detection methods are limited to cervical swabs performed in centralized laboratory settings, which require trained personnel, and are not feasible for in-home self-testing or mass screening in resource-constrained and conservative societies. Therefore, there is an urgent need for simple, accurate, specific, and user-friendly detection platforms to enable early diagnosis of HPV and related cancers.

Recently, clustered regularly interspaced short palindromic repeats (CRISPR)/CRISPR-associated nuclease (Cas) systems have been demonstrated to be sensitive, specific, and programmable for point-of-care diagnostics of infectious viruses. The CRISPR/Cas system, originally designed for genome editing, comprises two main components: a guide RNA (gRNA) and a Cas nuclease, forming a ribonucleoprotein (RNP). The gRNA identifies a complementary target, and the Cas nuclease cleaves that target (*cis*-cleavage)^14,15^. However, Cas12 and Cas13 nucleases exhibit collateral cleavage (*trans*-cleavage) of bystander DNA and RNA molecules^16,17^. The collateral cleavage feature of Cas12 and Cas13 proteins was successfully combined with quenched fluorophore DNA or RNA reporters: upon recognizing the target pathogen DNA or RNA sequence, the activated Cas effector degrades the reporter and releases the fluorophore. The collateral cleavage activity of the Cas effector was initially applied to precisely detect the Zika virus. The sensitivity of the CRISPR-Cas diagnostic platform was enhanced through target preamplification via isothermal amplification through Recombinase Polymerase Amplification (RPA) or Loop-Mediated Isothermal Amplification (LAMP). Multiple CRISPR-Cas system modalities using RPA or LAMP, known as SHERLOCK, SHERLOCK-like, HOLMES, HOLMESv2, and ISCAN, achieved clinically relevant sensitivity and specificity^17–20^. CRISPR-based diagnostics were comprehensively reviewed previously^21^. In these systems, nucleic acids are pre-amplified via polymerase chain reaction (PCR) or isothermal amplification^22^. PCR boasts high specificity and sensitivity but is centralized, requiring costly equipment, trained personnel, and long turnaround times^23^. Isothermal amplification methods offer a decentralized, cost-effective alternative but may amplify non-target nucleic acids^24,25^.

Combining the sensitivity of isothermal amplification and the specificity of CRISPR/Cas holds the potential to establish a new decentralized gold-standard point-of-care diagnostic test. Such platforms potentially meet the WHO ASSURED (affordable, sensitive, specific, user-friendly, rapid, equipment-free, delivered)^26^ guidelines for point-of-care diagnostics development. However, the existing CRISPR-based detection platforms are not suitable for multiplex detection of hrHPVs in a one-pot assay, primarily due to reaction chemistry incompatibility and nonspecific cross-collateral cleavage activity of the reporter molecules by different Cas effectors, necessitating separate reactions for different targets or controls. Similarly, multiple-target detection via LAMP-based amplification has posed challenges related to specificity^27–29^. To overcome these issues and create a highly sensitive and specific detection system, we previously coupled LAMP amplification with CRISPR detection in a one-pot setup for infectious virus nucleic acid detection^30^. However, detecting more than two targets remained challenging due to the lack of thermostable Cas effectors compatible with LAMP. Recent developments, including the discovery of thermostable Cas13 effectors like TccCas13a^30^ and HheCas13a^31^, along with multiple thermostable Cas12 effectors like AapCas12b^20^ and BrCas12b^32^, have expanded the multiplex detection possibilities. One-pot multiplexed detection of cancer-causing hrHPVs would enable early treatment of cervical cancers, intervention strategies, and policy development, potentially decreasing the toll of cervical cancer.

Here, we designed, built, and tested the CRISPRD platform, a novel approach capable of simultaneously and sensitively detecting three independent nucleic acid targets in a one-step, one-pot reaction in less than one hour. The CRISPRD platform employs a two-layer amplification system to maximize sensitivity (limit of detection, LoD, of 10 copies) and demonstrates high specificity for hrHPV subtypes 16 and 18, and the internal positive control RNase P. There is a critical need for hrHPV screening in both males and females, and CRISPRD seeks to fulfill this unmet need. The one-pot CRISPRD point-of-care diagnostic technology has the potential to address the global demand for hrHPV detection tests and can be adapted for self-testing or at-home hrHPV tests. We believe that this groundbreaking CRISPR-based diagnostic technology will bridge the gap and offer a practical point-of-care solution.

## Results and Discussion

### The CRISPRD workflow

We aimed to create a user-friendly one-pot multiplex diagnostic assay (<u>CRISPR</u>-based multiplexed <u>d</u>iagnostic test (CRISPRD) for the rapid mass screening of hrHPV subtypes. CRISPRD uses multiple layers of nucleic acid isothermal amplification to achieve high-level sensitivity and three different thermostable Cas effectors for specifically detecting three different targets. The initial step in the CRISPRD workflow involves a LAMP reaction to amplify the target DNAs at a constant temperature. The amplicons generated by LAMP are used as input for the subsequent detection either directly by Cas12b or indirectly by Cas13 variants (detecting RNA transcripts—all in one pot) (Figure 1).

**Fig. 1:**
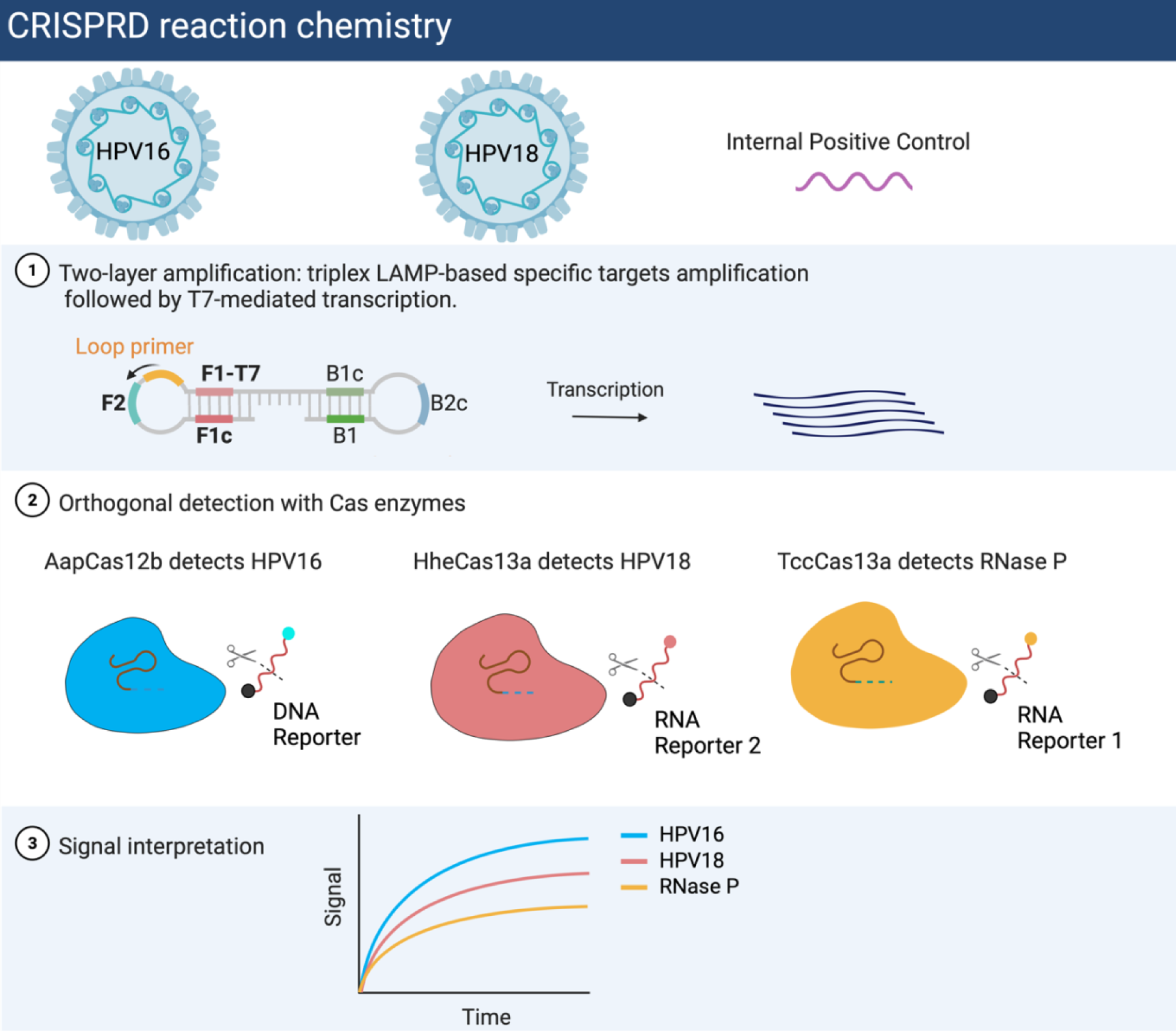
CRISPRD workflow for hrHPVs multiplex detection. **a.** CRISPRD is designed to detect three targets (HPV16, HPV16, and internal control) simultaneously. The two layers of amplification, LAMP for DNA and T7-transcription for RNA are followed by CRISPR-based specific detection of the three targets in one pot. CRISPRD deploys three orthogonal CRISPR Cas effectors (TccCas13a, HheCas13a, and AapCas12b) to detect its cognate target and digest its associated DNA or RNA reporter only. Target-induced Cas variant-based digestion of the reporter releases the fluorescence as a visual readout of pathogen detection.

We previously developed a one-pot reaction chemistry that couples LAMP amplification, T7-based transcription, and Cas13-based detection to efficiently identify infectious pathogens^30^. T7-based transcription was introduced in this modality to make RNA for Cas13-based detection. Here, we aimed to optimize the one-pot detection chemistry for multiplex detection. We designed several specific primer sets with PrimerExplorer V5 (https://primerexplorer.jp/e/) to specifically amplify three targets: HPV16, HPV18, and the human internal control (RNase P). To ensure efficient RNA transcription, we introduced a T7 promoter in the LAMP primers. The amplified target DNA activates Cas12b and initiates the promiscuous cleavage of DNA reporters; whereas the RNA resulting from transcription of the target activates Cas13 and initiates the promiscuous cleavage of RNA reporters. In both cases, reporter cleavage releases a distinct fluorophore and measurement of the resulting fluorescent signal is used for target detection. Next, we detected individual targets in single-pot reactions to ensure that the operating temperature and the reaction buffer were compatible with all enzymes.

For successful multiplexing detection, all system components must be orthogonal (only reacting with a cognate partner with no interference with other components). For that reason, orthogonality was assessed on three metrics: 1) reporter cleavage preference, i.e., finding individual Cas effectors that cut only one distinct reporter; 2) orthogonal fluorescence with the goal of choosing fluorophores with distinct excitation–emission spectra; and 3) orthogonal gRNA/Cas activation with the goal of finding Cas effectors activated by their distinct gRNA only. For multiplex diagnostics, we chose two Cas13a effectors (TccCas13a and HheCas13a), each preferring a specific RNA reporter for *trans* cleavage^30,31^ and AapCas12b, which is widely known to cut single-stranded DNA (ssDNA) reporters. Similarly, we designed three reporters each with different fluorophores, FAM, HEX, and ROX (one specific for each Cas effector) having different excitation–emission peaks, to detect the three independent targets.

Importantly, the entire CRISPRD platform operates within a closed-lid single-pot reaction to prevent cross-contamination. Additionally, the two steps, nucleic acid (DNA and RNA) amplification and gRNA–Cas effector-based detection operate at a single temperature, eliminating the requirement for a thermal cycler and ensuring the feasibility of CRISPRD for point-of-care use. Our hypothesized triplex platform is suited for detecting cervical cancer-causing HPV16 and HPV18 along with an internal control in a one-pot reaction at a single temperature. CRISPRD can potentially decentralize cervical cancer screening programs and enable cost-effective and convenient home testing.

### Thermophilic Cas effectors successfully detected individual targets

Although CRISPR-based diagnostics emerged as a promising point-of-care diagnostic system, it faces limitations in simultaneously detecting multiple targets lack of orthogonality among different Cas effectors, and cross degradation of the reporters by the multiple Cas effectors used. The emergence of thermostable Cas effectors may increase the multiplexing potential of CRISPR-Dx. For example, HheCas13a recognizes RNA targets and preferentially digests poly U-containing RNA reporters, while recently discovered thermostable TccCas13a recognizes RNA targets and preferentially cuts rArG-residue-containing RNA reporters. Based on the fact that Cas12 cuts DNA not RNA, we hypothesized that thermostable Cas effectors TccCas13, HheCas13a, and AapCas12b could be efficiently coupled with LAMP-based amplification to detect multiple hrHPVs (HPV16 and HPV18) and an internal control gene (human RNase P) in a one-pot reaction (Figure 2a).

**Fig. 2:**
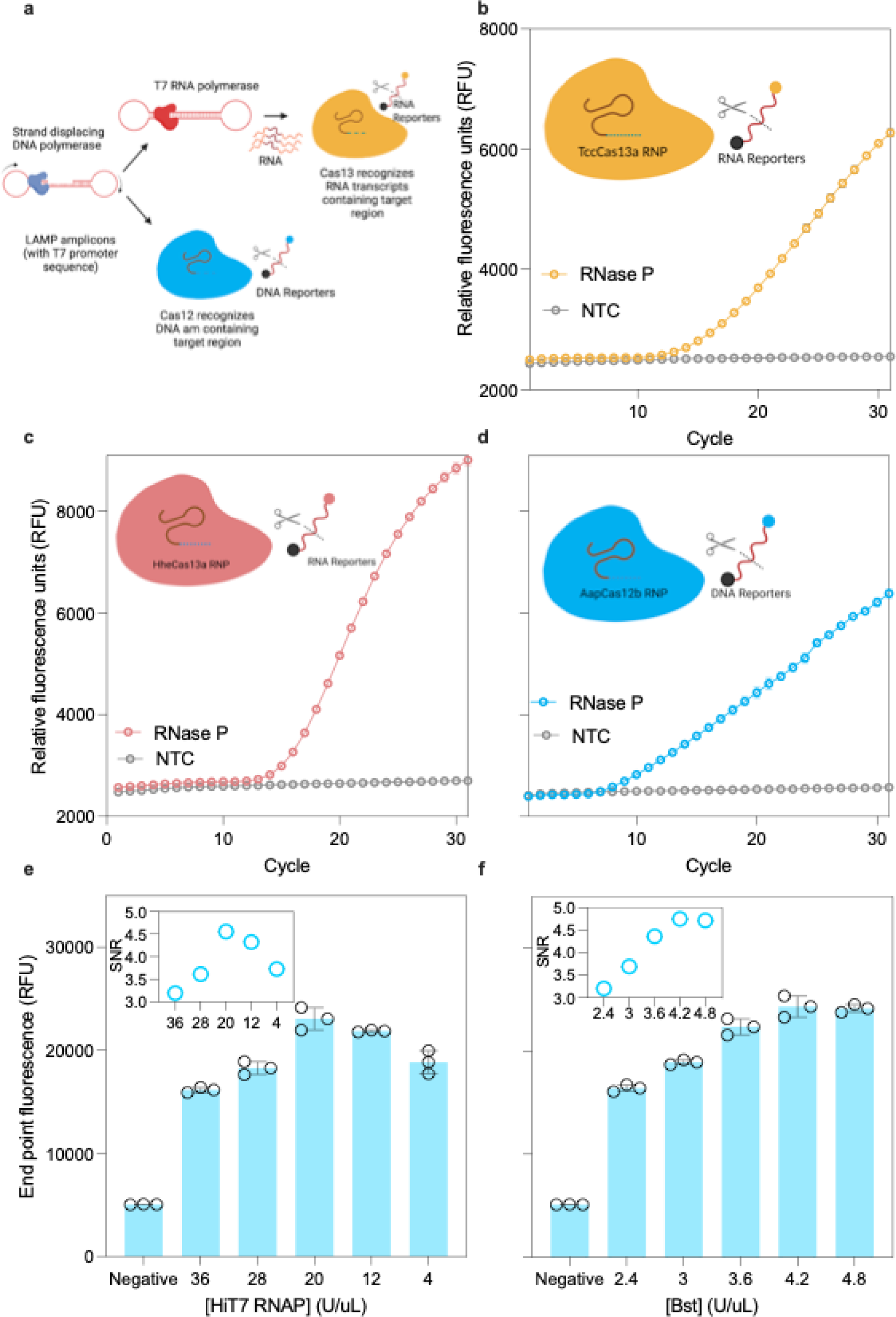
Optimization of CRISPRD single target reaction chemistry. **a.** Schematic of the single target detection. Individual target is amplified by LAMP and either detected directly by Cas12b or transcribed to RNA then detected by TccCas13a and HheCas13a **b-d.** Testing the CRISPRD reaction for the detection of RNase P synthetic DNA (1 ng per reaction) with TccCas13a (**b**), HheCas13a (**c**), and AapCas12b (**d**). **e-f.** Optimizing the CRISPRD detection chemistry by optimizing the concentration of HiT7 RNAP (e) and Bst DNA polymerase (f). The signal-to-noise ratio, calculated by dividing the signal over the background (reactions lacking target), is overlayed in both figures. SNR: signal-to-noise, NTC: no template control, RNP: Ribonucleoprotein, cycle = 2 minutes. The unlabeled y-axis follows the labeling of the y-axis in the left figure. All reactions were done in three replicates and means and standard deviation were plotted.

To confirm our hypothesis, we purified thermostable Cas effectors TccCas13a, HheCas13a, and AapCas12b^30^ and tested them individually along with synthetic DNA (RNase P) and selected LAMP primer sets to detect each target. As shown in Figure 2b–d, RNase P was efficiently detected by all Cas effectors. Next, we optimized the concentration of Bst DNA polymerase and HiT7 RNA polymerase to improve the detection efficiency of the CRISPRD platform. Our results demonstrated that adjusting the polymerase concentration substantially improved the signal (Figure 2e–f). Our single-target detection results confirmed that CRISPRD can enable the point-of-care screening for HPV, and offer a streamlined opportunity for multiplexed one-pot detection of the hrHPVs that cause cervical cancer.

### CRISPRD efficiently detected hrHPV DNA

Upon persistent infection, hrHPV integrates part of its DNA genome into the host genome; therefore, it is necessary to design a diagnostic method that detects the hrHPV integrated sequence^33^. Accordingly, we selected the permanently integrated region encompassing the E6 and E7 genes of HPV16 and HPV18 for CRISPRD detection.

To establish our CRISPRD system for hrHPV detection, we designed several LAMP primer sets using Primer Explorer software (http://primerexplorer.jp/lampv5e/index.html). We designed multiple gRNAs specific to the E6 and E7 amplicons produced from each primer set for HPV16 and HPV18 and screened for the best-performing primer/gRNA set for each target (Fig. S1, S2). The primer set C and gRNA #99 combination was superior for detecting HPV16 (using the AapCas12b RNP), whereas the primer set F and gRNA #92 combination displayed the most optimal performance for HPV18 (using the HheCas13a RNP). To establish an internal positive control, we sought to detect the widely abundant RNase P gene in humans with TccCas13a using a previously developed primer set^34^.

The CRISPRD reaction must be sensitive and specific to only HPV16 or HPV18 and should not cross-react with the human genome. We assessed the LoD for detecting HPV16 and HPV18 as well as RNase P with various dilutions of synthetic DNA, ranging from 0 to 1 million copies per uL, we found that our CRISPRD reactions could detect as low as 10 copies/uL of HPV16, HPV18, and RNase P (equal to 250 copies/reaction), comparable to Cobas HPV test from Roche^35^ (Fig. 3a, 3d, 3g). Of note, in the case of HPV18, we began the study by designing four primer sets and set A was the best performing. However, our CRISPRD reactions using Set A for HPV18 could not detect below 10,000 copies/uL (Fig. S3). To overcome this, we designed multiple LAMP primer sets and screened them for a better-performing primer set for detecting HPV18. Eventually, primer set F with gRNA#92 demonstrated very sensitive detection of HPV18 (Fig. 3d). This indicates that robust primer screening can improve the sensitivity of a test quite significantly.

**Fig. 3:**
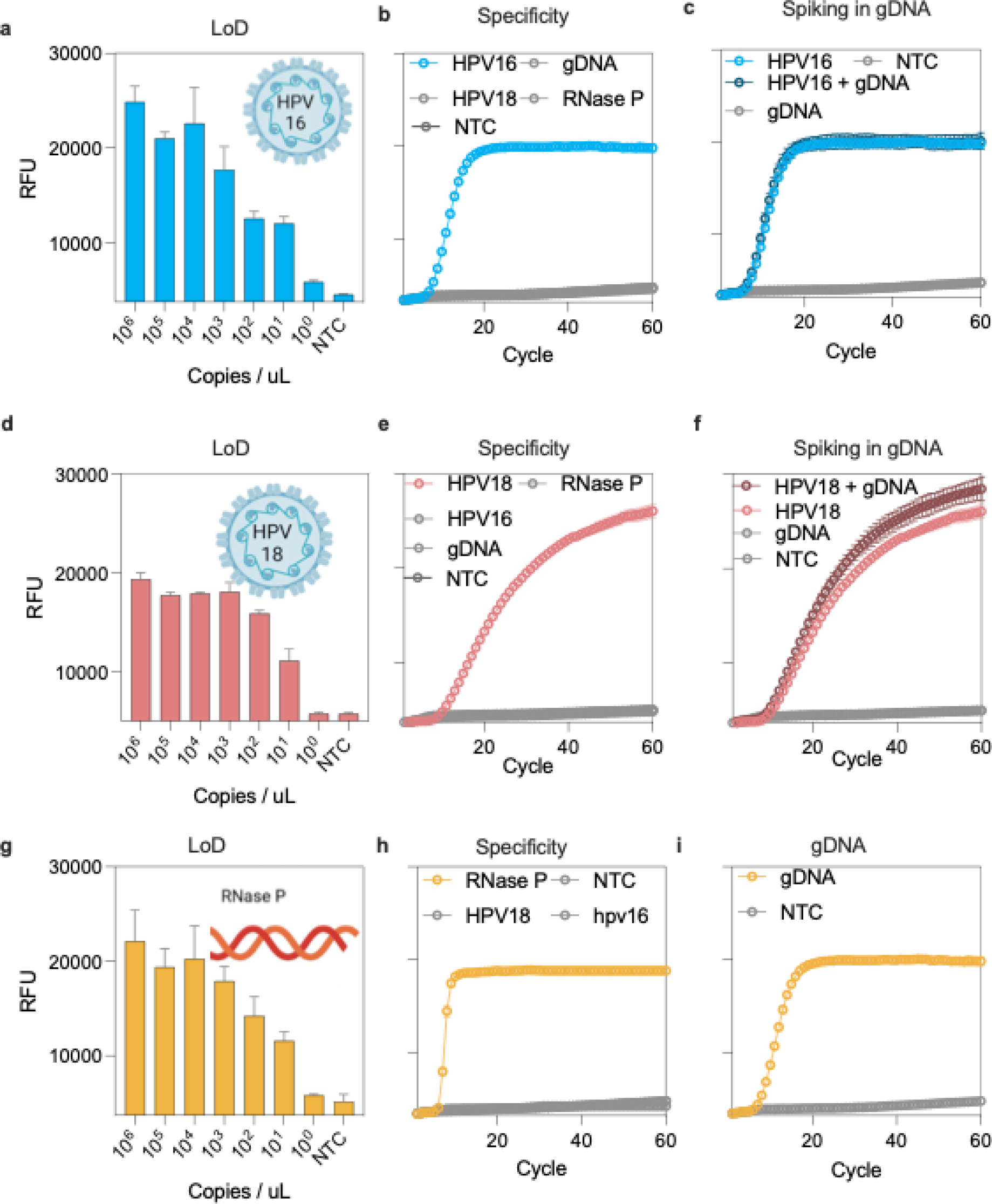
CRISPRD-based sensitive and specific detection of HPV16 and HPV18 synthetic DNA. **a.** LoD for HPV16 with primer set C and crRNA# 99 with AapCas12b. **b.** Specificity of the CRISPRD reaction for HPV16 detection. The reaction was incubated with 1 ng of either HPV16, HPV18, gDNA, or RNase P. **c.** Spiking in gDNA: HPV16 was spiked into 100 ng/uL of gDNA to a final concentration of 1 ng/uL. **d.** LoD for HPV18 using primer set F and crRNA #92. **e.** Specificity of the CRISPRD reaction for HPV18 detection. The reaction was incubated with 1 ng of either HPV16, HPV18, gDNA, or RNase P. **f.** Spiking in gDNA: HPV18 was spiked into 100 ng/uL of gDNA to a final concentration of 1 ng/uL. **g.** LoD for RNase P with primer set Pop7 and crRNA# 92 with TccCas13a. **b.** Specificity of the CRISPRD reaction for RNase P detection. The reaction was incubated with 1 ng of either HPV16, HPV18, or RNase P. **c.** detection of RNase P in gDNA: 10 ng of gDNA were incubated with CRISPRD reaction for RNase P detection. NTC: No template control, RFU: relative fluorescence units, Cycle: two minutes, all reactions were done in three replicates, and the mean and standard deviation were plotted.

Next, we conducted a specificity test by incubating human DNA in the CRISPRD reaction targeting HPV16, HPV18, or RNase P. The test exhibited specificity towards HPV16 and HPV18, showing no interference with human genomic DNA (Fig. 3b, e). Furthermore, CRISPRD successfully detected RNase P in human genomic DNA (Fig. 3i). Additionally, each gRNA-Cas specifically detected its respective target without cross-reactivity with other targets (Fig. 3b, e, and h). Moreover, we spiked HPV16 and HPV18 into human genomic DNA and confirmed that the CRISPRD method detected both HPV16 and HPV18 precisely (Fig. 3c, f, and i).

### Development of a CRISPRD one-pot multiplexed detection assay

#### Orthogonality assessment

Our next objective was to verify the complete orthogonality of the system in terms of 1) gRNA activation, 2) target detection, 3) fluorescence readout, and 4) reporter cleavage preference. To confirm that the gRNAs are not interchangeable between Cas effectors, we conducted reciprocal incubations of TccCas13a with the gRNAs for HheCas13a or AapCas12b (same setup for all Cas effectors) to ensure that gRNAs would solely activate their respective Cas13 effectors without any cross-reactivity. Our findings confirmed that the gRNAs reacted specifically with their cognate Cas effector (Fig. 4a). Further, we swapped RNPs with different reactions to confirm that each RNP is solely activated by its respective target. To this end, we incubated the TccCas13a RNP targeting HPV16 in a CRISPRD reaction to see if it was aberrantly activated by HPV18 or RNase P. We conducted similar experiments to test the target specificity of HheCas13a and AapCas12b. Our results demonstrated that the RNPs were activated only in the presence of their specific target (Fig. 4b).

**Fig. 4:**
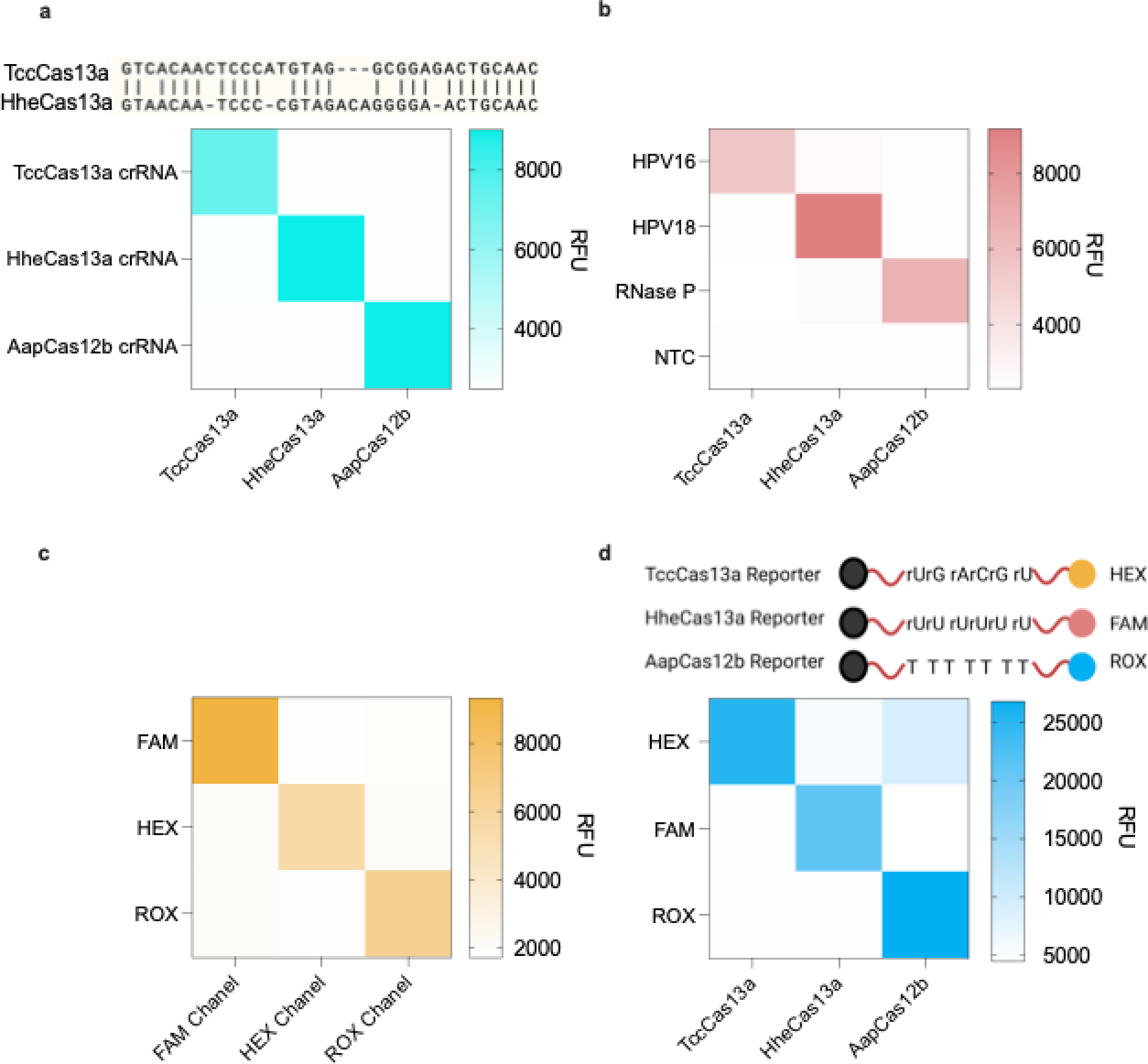
CRISPRD orthogonality. **a.** Orthogonality of CRISPR/Cas activation. Each Cas effector was allowed to form an RNP with other effector’s crRNAs and then proceeded to detect corresponding targets. **b.** Orthogonality and specificity of the target. Each RNP was incubated with all three different targets to test whether targets can activate incorrect RNPs. **c.** Orthogonality of fluorophores excitation and emission. Three different CRISPRD reactions were carried out to detect RNase P and cut reporters carrying three different fluorophores. Each reaction was read by three different channels in the qPCR machine (Bio-Rad). **d.** Orthogonality of reporter preference. Activated Cas effectors were incubated with different reporters to check if there was any cross-talk in signal reporting. For all reactions, we conducted testing on three replicates and visualized the mean signal on a heat map.

Next, we focused on establishing the orthogonality of the reporter systems. We determined that HEX, FAM, and ROX offered the highest sensitivity for detection (Fig. S4). We validated the orthogonality of these fluorophores by subjecting the channels of the fluorometer to all signals. The flourophores exhibited distinct emissions spectra. Specifically, the FAM channel exclusively detected the FAM reporter, the HEX channel exclusively detected the HEX reporter, and the ROX channel exclusively detected the ROX reporter (Fig. 4c).

Next, we incubated activated RNPs from each of the three effectors with each reporter to confirm that each Cas effector would only cleave the corresponding reporter and not interfere with other reporters in the reaction. We found that both of the Cas13 effectors cleaved only their corresponding reporter molecule (Fig. 4a). Nonetheless, TccCas13a exhibited slightly lower activity toward the Poly-U RNA reporter, but this effect was diminished by reducing the Poly-U reporter concentration (data not shown).

Initially, we did not plan to test the activity of AapCas12b (an RNA-guided DNA nuclease) on RNA reporters since it is widely established to cut only ssDNA by collateral cleavage. However, when we later duplexed TccCas13a and AapCas12b, to our surprise, We noticed a nonspecific signal and thought it was coming from contamination. (Fig. S5). We then ensured that the reagents were clean and re-tested, but observed the same nonspecific signal. We assumed that AapCas12b and TccCas13a were not compatible in duplex reactions. Recently, Dmytrenko et al. showed that Cas12a2 has collateral cleavage activity on ssDNA, dsDNA, and ssRNA^36^. We questioned whether AapCas12b has collateral cleavage activity on RNA, causing this nonspecific signal. Indeed, when we incubated an activated AapCas12b RNP with rArG reporters, a signal emerged indicating that the reporter was cleaved (Fig. S6a). We then incubated the AapCas12 RNP with different RNA reporters hoping that it might have a preference for certain RNA nucleotides, but it cleaved most RNA reporters (except poly-U reporters used with HheCas13a) (Fig. S7, S8). Then, we re-purified AapCas12b from different clones and confirmed the AapCas12b-based promiscuous degradation of RNA reporters (Fig. S6b).

To overcome the issue of aberrant cleavage by AapCas12b on RNA reporters, we purified another thermostable Cas12 effector. Nguyen et al. identified BrCas12b as a thermostable Cas12b with superior activity with LAMP compared to AapCas12b^32^. The group further engineered an enhanced version (eBrCas12b) that could operate at the optimal temperature range for LAMP (60– 65°C)^37^. We purified BrCas12b and eBrCas12b to test their activity on RNA reporters. However, similar to AapCas12b, BrCas12b and eBrCas12b also digested RNA reporters (Fig. S9), confirming that Cas12b effectors might share a universal RNA trans-cleavage capability.

We wanted to utilize this dual nuclease feature of AapCas12b using DNA and RNA reporters carrying the same fluorophore. We hypothesized that this dual nuclease feature might result in two cleavage events per collateral cleavage round, boosting the signal, speed, and sensitivity. However, the emergent signal from both reporters was lower than that of ssDNA reporters alone (Fig. S10).

The discovery that AapCas12b cleaves RNA might have implications for genome editing applications. The application of Cas12 in genome editing as an alternative to Cas9 was enabled by the fact that Cas12 has *trans*-cleavage activity on ssDNA, which is not abundant *in vivo*. But considering that AapCas12b has collateral cleavage activity on RNA, its applications in genome editing might be limited as editing events will be accompanied by promiscuous cleavage of bystander RNA, leading to uncontrollable outcomes.

### CRISPRD-based duplex detection of hrHPV

The duplexed CRISPRD system included standard reagents for each CRISPRD reaction, the components for duplexed LAMP, two assembled Cas RNPs, and two reporter molecules. We first tested the compatibility of TccCas13a and HheCas13a in our one-pot CRISPRD system (Fig. 5a) and found that these two Cas enzymes were compatible in the CRISPRD duplex reaction (Fig. 5b). Indeed, each effector was activated and cleaved its respective reporter only when the appropriate target was present. We obtained similar results with HheCas13a and AapCas12b (Fig. 5c–d).

**Fig. 5:**
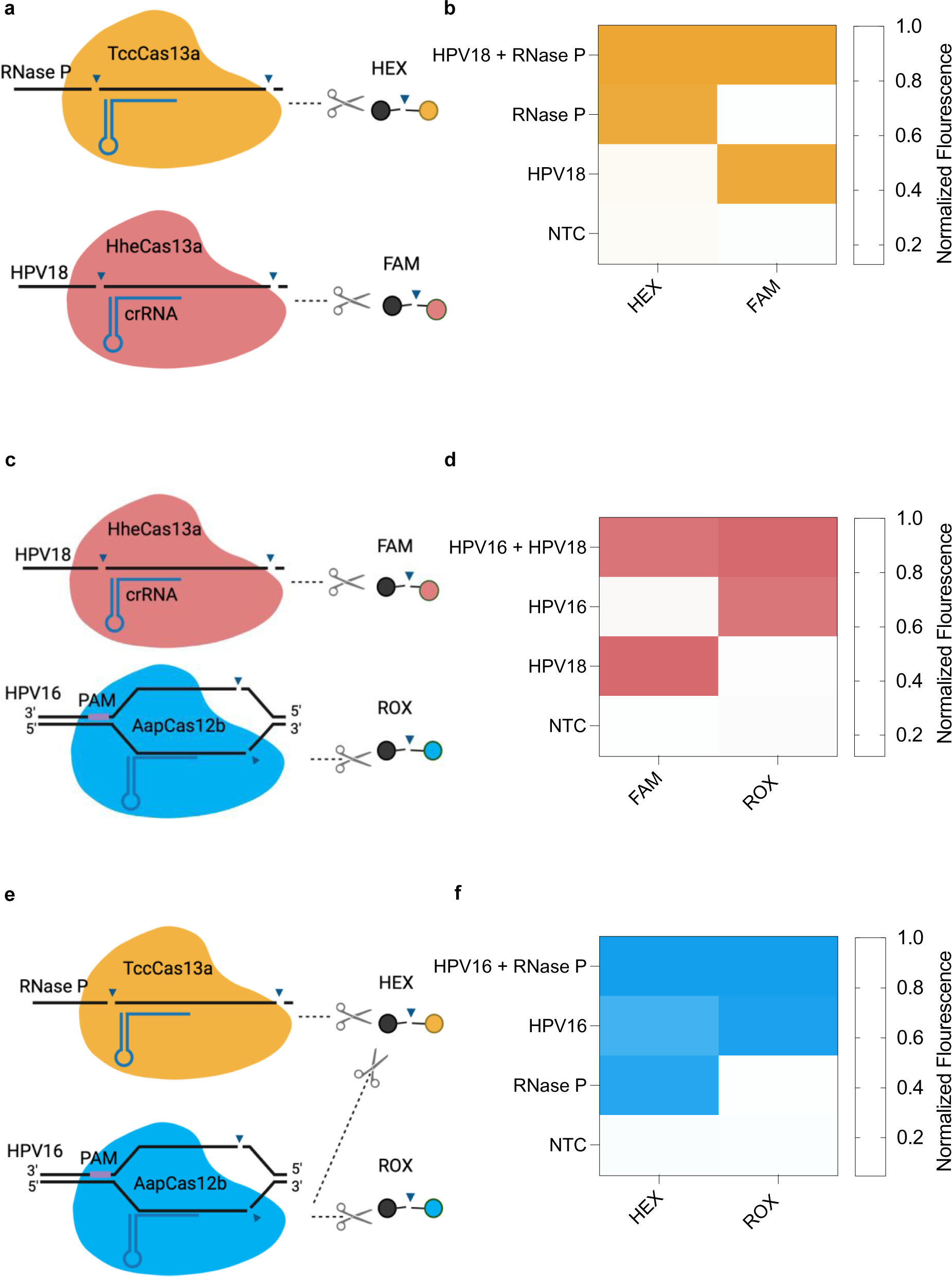
Proof of concept for duplex detection with TccCas13a, HheCas13a, and/or AapCas12b. **a.** schematic of the duplexing CRISPRD reaction using HheCas13a (targeting HPV18 and trans-cleaving FAM reporters) and TccCas13a (targeting RNase P and trans-cleaving HEX reporters). **b.** Duplexing results with HheCas13a and AapCas12b. **c.** schematic of the duplexing CRISPRD reaction using AapCas12b (targeting HPV16 and trans-cleaving ROX reporters) and TccCas13a (targeting RNase P and trans-cleaving HEX reporters) **d.** Duplexing results with HheCas13a and TccCas13a. **e.** schematic of the duplexing CRISPRD reaction using HheCas13a (targeting HPV18 and trans-cleaving FAM reporters) and AapCas12b (targeting HPV16 and trans-cleaving ROX reporters). **f.** Duplexing results with HheCas13a and AapCas12b. Normalized fluorescence was calculated by dividing the signal over the highest signal from the corresponding reporter (n = 3; the mean of the normalized fluorescence is displayed on the heatmap).

While both AapCas12b and TccCas13a possess the ability to cleave rArG reporter molecules (Fig. 4a), only AapCas12b can effectively cleave poly-T reporters, facilitating the identification of the activated enzyme (Fig. 5e). Activation of AapCas12b results in the cleavage of both poly-T and rArG reporters, resulting in ROX and HEX fluorescence. Conversely, activation of TccCas13a leads to the cleavage of solely rArG reporters, resulting only in HEX fluorescence. We designed TccCas13a for detecting the internal control RNase P and AapCas12b for detecting HPV16. HPV16-positive samples exhibit a signal in both the ROX and HEX channels, while negative samples display a signal only in the HEX channel. The assay is considered invalid if no signals are observed in either channel, indicating an unreliable sample.

### Development of one-pot CRISPRD for multiplexed HPV16, HPV18, and RNase P detection

We then sought to detect three targets in a single-tube multiplexed reaction. The triplex CRISPRD reaction contained the optimized reagents in addition to three LAMP primer sets, three Cas variant RNPs, and three reporters. Given the promiscuity of AapCas12b in cutting rArG reporters, we addressed this limitation by rearranging the targets (Fig. 6a–b): HheCas13a targets HPV18, AapCas12b targets HPV16, and TccCas13a targets RNase P.

**Fig. 6:**
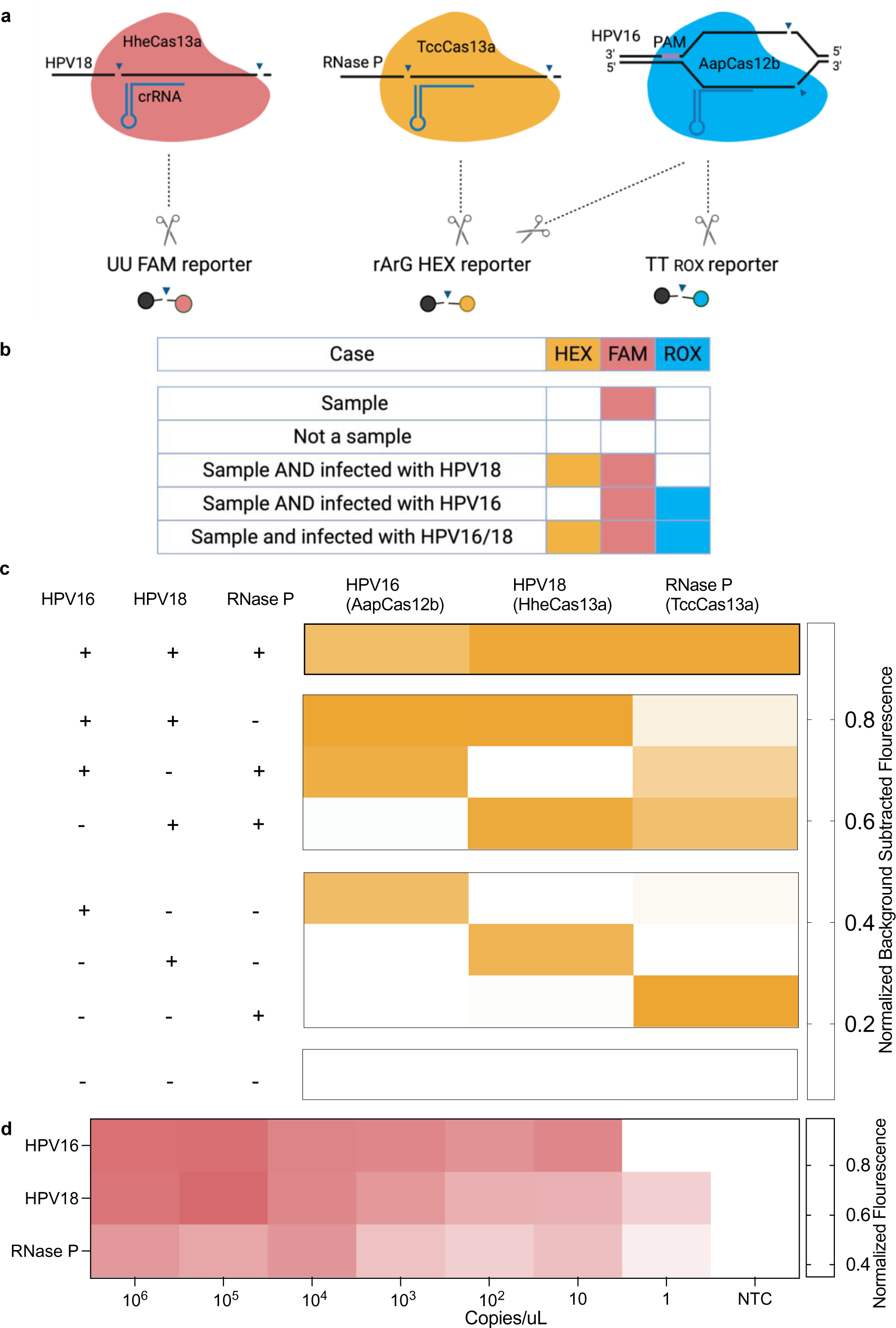
CRISPRD-based multiplex detection of HPV16, HPV18, and internal control. **a.** schematic of the triplexing CRISPRD reaction using HheCas13a (targeting HPV18 and trans-cleaving FAM reporters), TccCas13a (targeting RNase P and trans-cleaving HEX reporters) and AapCas12b (targeting HPV16 and trans-cleaving ROX reporters). **b.** Scenarios that prove that the cross-reactivity of AapCas12b with TccCas13a reporters will not affect the practical application**. c.** Proof of concept of the triplexing reaction. One master mix containing all reagents for a triplexing reaction was prepared without the addition of targets. Targets were added in a final 1 ng/uL to each tube according the to experimental design (Left), and the reaction proceeded for one hour **d.** LoD of the triplexing reaction. One master mix containing all triplexing reagents was prepared. Three targets were mixed in one tube from which multiple serial dilutions were made. The reaction proceeded for one hour. n=3; Normalized background-subtracted fluorescence: background fluorescence (from tubes containing no target) was subtracted from the fluorescence signal (tubes containing targets) and then normalized to the highest signal in the corresponding reporter.

In practice, the third target is always a positive internal control which indicates that the sample is intact. Accordingly, five different scenarios could result from this triplex reaction (Fig. 6b): 1) If a sample contains a sufficient amount of DNA from the test subject, it must contain the internal control, in this case RNase P, which would lead to the activation of TccCas13a and the cleavage of rArG HEX reporters; 2) If a sample is degraded or not enough DNA was collected, no HEX signal would emerge; 3) If the subject is infected with HPV16, this would lead to the activation of AapCas12b, which would cleave Poly-T ROX reporters and rArG HEX reporters. Theoretically, the presence of both signals would indicate the presence of HPV16 in the sample, but would not ensure the presence of RNase P. For diagnostic purposes, however, the presence of HPV16 is sufficient to indicate an intact sample; 4) If the subject is infected with HPV18, this would indicate it also contains RNase P, leading to the activation of HheCas13a and TccCas13a and the cleavage of Poly-U FAM and rArG HEX reporters, respectively; 5) A subject that is infected with HPV16 and HPV18 would have the three signals: HEX, ROX, and FAM.

For a proof of concept, we made all combinations of targets in a one-pot triplexed CRISPRD reaction (Fig. 6c). In one setup, we added all three targets to the reaction tube to ensure that triplexing is possible. Additionally, we added all two-target combinations in a triplexing reaction, and the signal that emerged was specific, except (as expected) in the combination of RNase P and HPV16 (due to the promiscuity of AapCas12b). We also added each target individually to the triplex CRISPRD reaction and detected each target in the corresponding channel for each single-target reaction.

To ensure that detecting three targets in one pot does not compromise the detection sensitivity, we assessed the LoD of each target by incubating serial dilutions of the targets in the CRISPRD reaction and attained a LoD of 10 copies/uL for all targets, and reached single-copy detection for HPV18 and RNase P (Fig. 6d).

### CRISPRD successfully detected HPV16 and HPV18 DNA in human cell lines

hrHPVs integrate part of their DNA genome into the host’s genome, and persistent expression of the virus oncogenes E6 and E7 leads to cervical malignancies. Human cell lines CaSki (HPV16) and HeLa (HPV18) integrated with HPV genomes were previously isolated and represent a good resource for hrHPV research^38,39^. Thus, we designed our CRISPRD method against the hrHPV-integrated sequence to confirm the efficacy of our diagnostic module and ensure it can detect HPV DNA integrated in human DNA. Following purification of gDNA from cell lines (Fig 7a), CRISPRD detected HPV16 and HPV18 in CaSki and HeLa cells, respectively, confirming the specificity and multiplexing potential of the CRISPRD reaction chemistry (Fig. 7b).

**Fig. 7:**
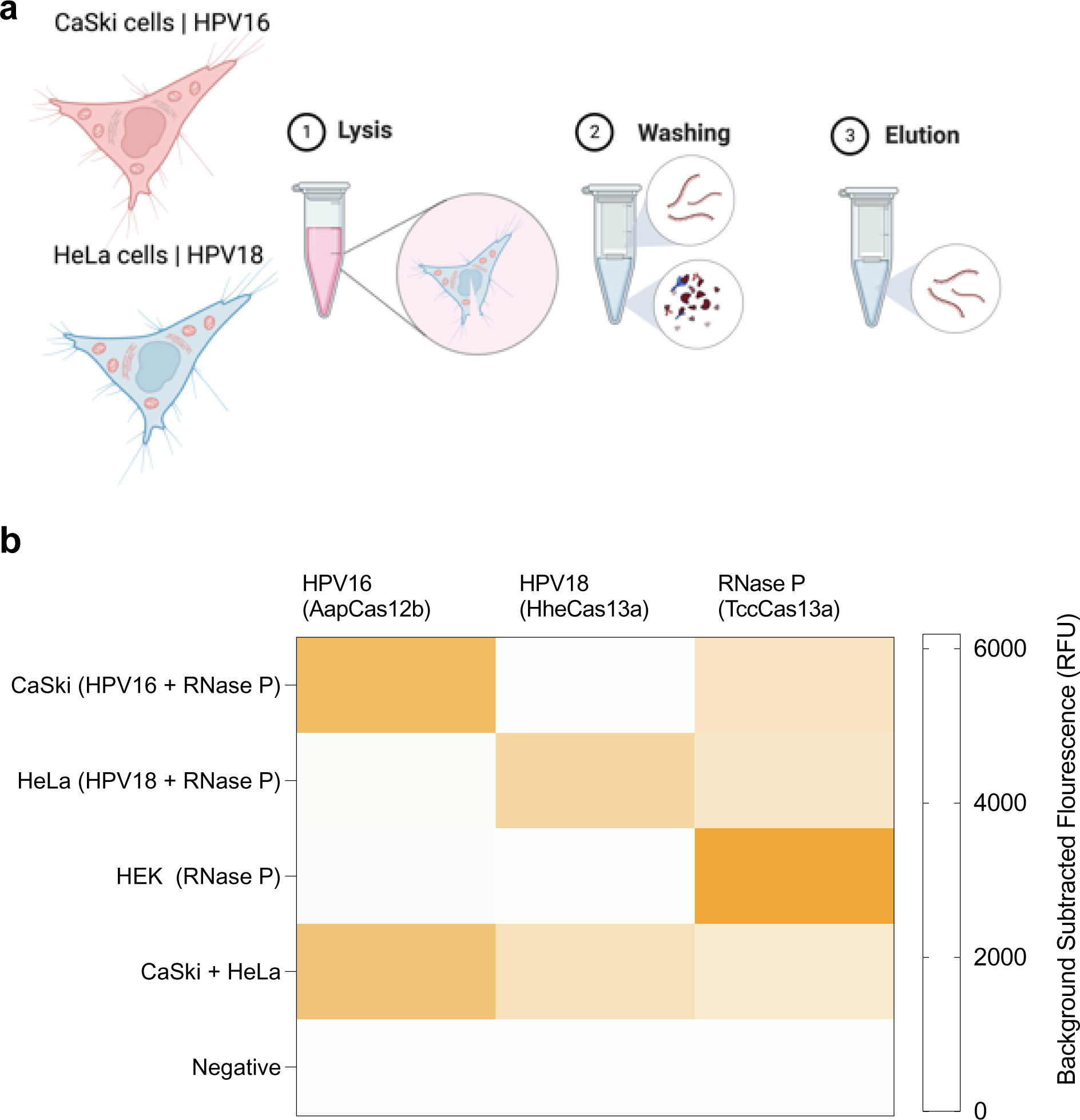
Validation of CRISPRD for detection of HPV DNA in cell lines. **a.** CaSki cells with integrated HPV16 genome and HeLa cells with integrated HPV18 genome were used to validate the triplex CRISPRD reaction. The genomic DNA was extracted by a standard protocol of lysis, washing, and elution. **b.** Validation of the CRISPRD reaction with cell lines. One master mix containing all reagents for a triplexing reaction was prepared without the addition of targets. Equimolar amount of each cell gDNA was mixed for the multiplexed detection of HPV16 and HPV18 gDNA targets in one pot. The reaction proceeded for two hours (n=3). HEK-293T cells were used as a negative control for HPV16 and HPV18. The mean of the background-subtracted fluorescence was visualized in a heatmap.

To date, most CRISPR-based platforms offer single-target detection in one pot. Multiplexing is challenging due to the nonspecific cleavage activities of Cas effectors. Arrayed multiplexing, exemplified by CARMEN^40^ and mCARMEN^41^, is one way to overcome the nonspecific cleavage activity of Cas effectors by performing multiple single-target detection events in distinct channels. Arrayed multiplexing enabled high-throughput multiplexing of 1000 targets in a single test^40^. However, arrayed multiplexing suffers from several limitations, including 1) the assay is not user-friendly and not compatible with point-of-care detection, 2) requires special, separate equipment for amplification and detection, and 3) is time-consuming: results take more than a day to obtain.

Single-tube multiplexing, on the other hand, was enabled by the discovery of new Cas effectors. The fact that Cas12 nucleases cut DNA whereas Cas13 nucleases cut RNA enabled duplex reactions. The discovery of collateral cleavage preference toward some nucleotides enabled multiplexing several Cas13 effectors, but detection was separated from amplification^42^. Recently, SHINE.V2 reported duplex detection of SARS-CoV-2 and an internal control using Cas13 and Cas12 in one pot (with amplification via RPA)^43^.

The CRISPRD platform can be scaled to many other diseases that require single, double, or triple biomarker detection. During the COVID-19 pandemic, there was a need for a test that differentiates COVID from Influenza to contain the spread of SARS-CoV-2 and influenza subtypes A and B. Our CRISPRD platform can be reprogrammed to detect COVID and subtype Influenza A and B. CRISPRD can also be applied to detect the deadly methicillin-resistant *Staphylococcus aureus* (MRSA), which requires detecting two genes (and a third for more confirmation)^27,44^.

Although the CRISPRD platform is a step forward toward the multiplex detection of pathogens at the point-of-care, CRISPRD has the same limitations of most CRISPR-based detection platforms. We previously outlined the challenges and potential solutions to bring CRISPR-based detection methods to the market^21^. Namely, the CRISPRD platform relies on fluorimetric reading of the signal. Though colorimetric signals are preferred, especially in point-of-care setups, companies such as Egoo Health are trying to commercialize cheap fluorescence readers that can read multiple fluorometric signals at once. Additionally, the CRISPRD platform still needs to be tested with quick sample processing modalities. An ideal diagnostic kit would have a sample-to-answer either in a single tube, or in multiple channels controlled by automation in small point-of-care devices^45^. An ideal diagnostic tool should also be straightforward, inexpensive, and sensitive^46^.

### Conclusion

In this study, we screened and coupled recently discovered thermostable Cas effectors with LAMP-based amplification to create a CRISPR-based one-pot platform called CRISPRD for multiplex detection of hrHPVs. CRISPRD enabled the first highly specific and sensitive triplex hrHPV detection method, a step forward to decentralizing hrHPV diagnostics and assisting in cervical cancer mitigation according to WHO recommendations. The one-pot, single-step reaction chemistry of the CRISPRD detection module fulfills the WHO-recommended ASSURED criteria of PoC detection and can be adapted for self-testing for hrHPV. Our CRISPRD platform offers unprecedented specificity relative to any multiplexed LAMP platform designed to date^27–29^. These advancements are crucial in mitigating HPV-related diseases, potentially saving millions of lives and curbing cervical cancer-related human suffering and economic losses. We anticipate the CRISPRD platform to be adapted in lab-free point-of-care environments as a user-friendly, low-cost, rapid, sensitive, easy-to-use diagnostic tool, filling the gap in hrHPV detection and assisting in early mitigation of cervical cancer.

## Materials and Methods

### Protein purification

The plasmid p2CT-His-MBP-Hhe_Cas13a_WT used for HheCas13a expression was acquired from Addgene (plasmid #91871). Recombinant HheCas13a was purified according to a previously published protocol^31^. The clone containing TccCas13a is also available in Addgene (Plasmid # 199754). Expression and purification of TccCas13a followed a previously published protocol^30^. The plasmid pAG001-His6-TwinStrep-SUMO-AapCas12b for expressing AapCas12b was obtained from Addgene (plasmid #153162), and purification followed a previously published protocol^47^.

Clones containing BrCas12b^32^ and eBrCas12b^37^ were obtained from Addgene (Plasmids #182276 and #195339, respectively). Expression and purification of BrCas12b and eBrCas12b were performed according to previously published protocols^32,37^. After cleavage with TEV protease (from Tobacco Etch Virus), size-exclusion chromatography was performed to separate BrCas12b or eBrCas12b from the MBP (Maltose Binding Protein) tag. The size exclusion buffer contained 25 mM Tris HCl pH 7.5, 100 mM NaCl, 10% glycerol, and 1 mM TCEP (tris(2-carboxyethyl) phosphine).

### gRNA production

To generate the HheCas13a and TccCas13a gRNAs, 10 uM of T7 promoter (oligo from IDT) was annealed to the bottom strand which contains the repeat region and the spacer for the gRNA (single-stranded oligo). Annealing occurred in a PCR buffer with a gradual decrease of temperature from 95 to 25°C, in 5°C decrements every two minutes. The annealed product was in vitro transcribed (IVT) with T7 highscribe kit (NEB) according to the manufacturer’s protocol. The RNA product was purified with zymo RNA purification kit, and measured with a nanodrop spectrophotometer.

To generate the AapCas12b, BrCas12b, and eBrCas12b gRNAs, the repeat region was used as a single-stranded DNA ultramer (IDT). To make a full-length DNA for the gRNA (containing the repeat and the spacer), the spacer was used as a reverse primer (having a shared region with the repeat region) and PCR amplified with the forward primer T7 oligo. The PCR product was purified with Qiagen and the product was measured with a nanodrop spectrophotometer. Then, 300–1000 ng of the PCR product was used as a template for the IVT using Hiscribe kit (NEB).

The product was purified with zymo RNA purification kit and measured with a nanodrop spectrophotometer.

### Design and screening of LAMP primers

Different primer sets targeting several regions of the E6 and E7 genes of the HPV16 and HPV18 genomes were designed using PrimerExplorer v5 software (https://primerexplorer.jp/e/). BLAST analysis of the designed primers showed their specificity to HPV16 and HPV18 genomes. Optimal primer sets that showed the best performance were determined by conducting colorimetric LAMP assays and CRISPRD detection assays to detect specific targets.

### CRISPRD reaction for single-, double-, and triple-target detection

The CRISPRD reaction master mix was created in two parts: RNP assembly (incubating Cas effector with gRNA (Table 1)) followed by making the master mix (Table 2). The tables below describe the reagents used in the CRISPRD single-, double-, and triple-target detection reactions.

**Table 1:**
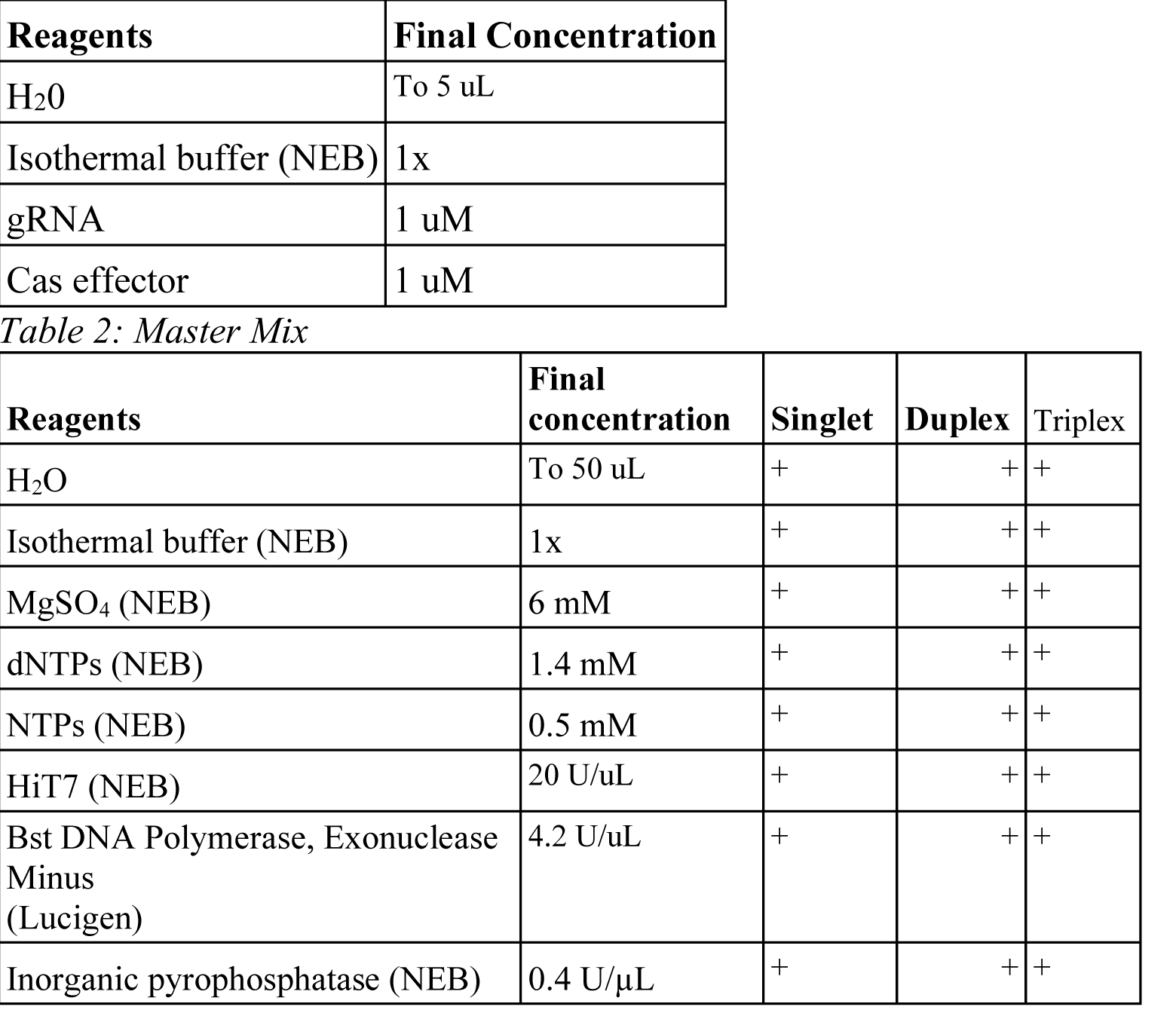
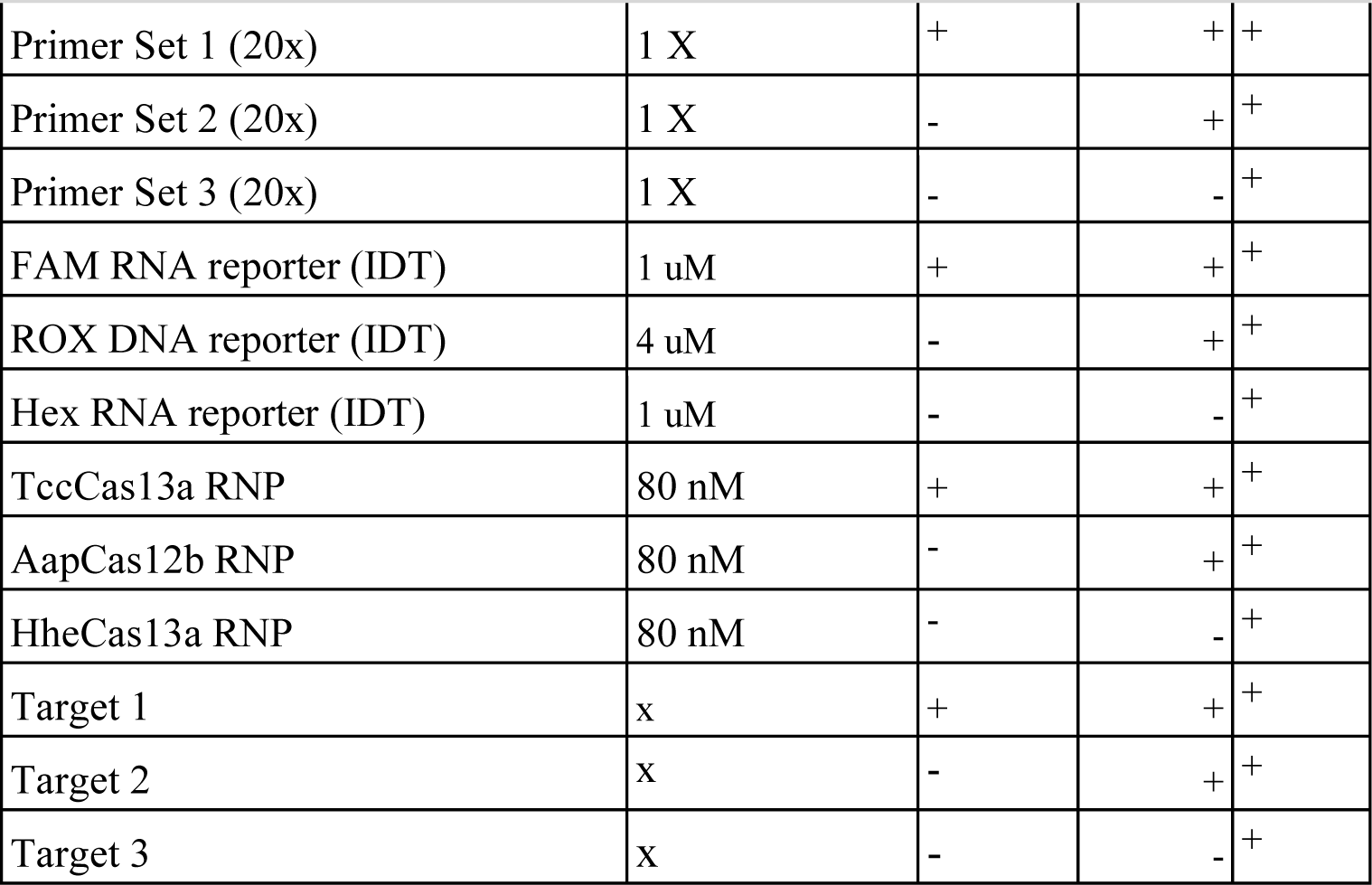
RNP assembly.

## AUTHOR INFORMATION

Corresponding Author: Magdy M. Mahfouz − Laboratory for Genome Engineering and Synthetic Biology, Division of Biological Sciences, 4700 King Abdullah University of Science and Technology, Thuwal 23955-6900, Saudi Arabia; orcid.org/0000-0002-0616-6365;

Authors

Ahmed Ghouneimy − Laboratory for Genome Engineering and Synthetic Biology, Division of Biological Sciences, 4700 King Abdullah University of Science and Technology, Thuwal 23955-6900, Saudi Arabia; https://orcid.org/0000-0003-3207-5181

Zahir Ali − Laboratory for Genome Engineering and Synthetic Biology, Division of Biological Sciences, 4700 King Abdullah University of Science and Technology, Thuwal 23955-6900, Saudi Arabia, https://orcid.org/0000-0002-7814-8908

Rashid Aman − Laboratory for Genome Engineering and Synthetic Biology, Division of Biological Sciences, 4700 King Abdullah University of Science and Technology, Thuwal 23955-6900, Saudi Arabia; https://orcid.org/0000-0002-6549-4887

Wenjun Jiang, − Laboratory for Genome Engineering and Synthetic Biology, Division of Biological Sciences, 4700 King Abdullah University of Science and Technology, Thuwal 23955-6900, Saudi Arabia.

Mustapha Aouida − Division of Biological and Biomedical Sciences, College of Health and Life Sciences, Hamad Bin Khalifa University, Education City, Qatar Foundation, Doha, Qatar, P.O. Box: 34110).

## Author Contributions

MM conceived the research. AG, ZA, and RA designed the research. AG, ZA, RA, and WJ performed the research. MA contributed reagents including cell lines. MM, AG, and ZA wrote the paper with input from RA and WJ.

## Notes

The authors declare competing financial interests.

## ASSOCIATED CONTENT

Supporting information 1

Supplementary Figure 1. | Screening for the best-performing set of primers/crRNAs (real-time data).

Supplementary Figure 2. Primer screening for HPV16 and HPV18 (Endpoint). Supplementary Figure 3. LoD using primer set <u>A</u> for HPV18.

Supplementary Figure 4. Reporter screening.

Supplementary Figure 5. Duplexing AapCas12b with TccCas13a.

Supplementary Figure 6. Testing the collateral cleavage activity of AapCas12b purified from two different clones.

Supplementary Figure 7. Activity of AapCas12b on RNA reporters with different nucleotide compositions.

Supplementary Figure 8. Reporter screening with AapCas12b real-time data. Supplementary Figure 9. BrCas12b cleaves RNA and DNA reporters.

Supplementary Figure 10. Testing AapCas12b with RNA and DNA reporters.

Supplementary information 2. Nucleic acid sequences used in this study

## Data Availability

All data produced in the present study are available upon reasonable request to the authors

## Acknowledgments

We would like to thank genome engineering and synthetic biology laboratory members for insightful discussions and technical support.

## Funding

This work was supported, in part, by the Smart Health Initiative at KAUST to MM.

## Notes

### Competing Interest Statement

The authors have declared no competing interest.

