## Supplementary material for "CRISPR-based multiplex detection of human papillomaviruses for one-pot point-of-care diagnostics": NA

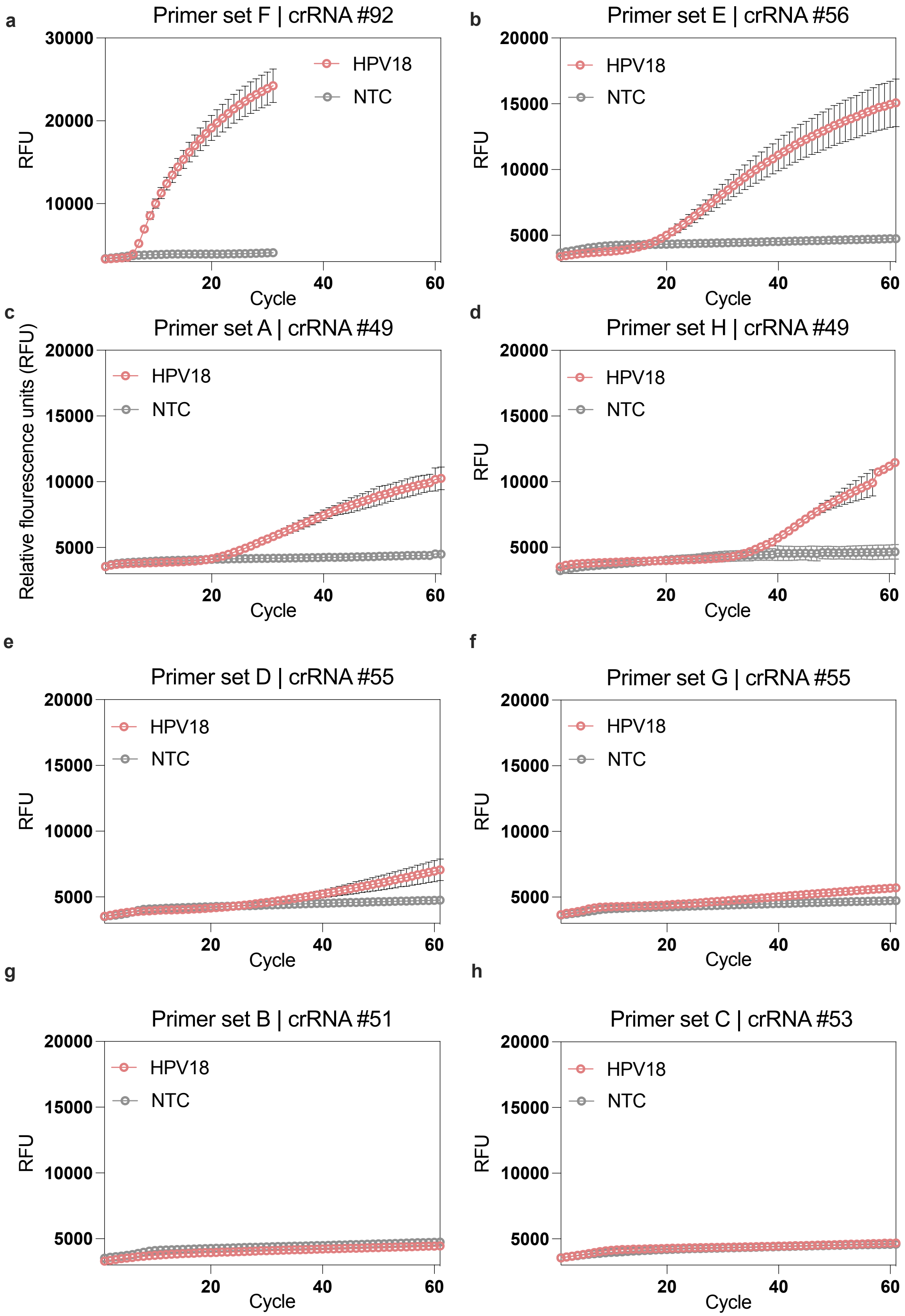
a
Primer set F | crRNA #92
RFU
30000
20000
10000
HPV18
NTC
20
40
60
Cycle
b
Primer set E | crRNA #56
RFU
20000
15000
10000
5000
HPV18
NTC
20
40
60
Cycle
c
Primer set A | crRNA #49
Relative flourescence units (RFU)
20000
15000
10000
5000
HPV18
NTC
20
40
60
Cycle
d
Primer set H | crRNA #49
RFU
20000
15000
10000
5000
HPV18
NTC
20
40
60
Cycle
e
Primer set D | crRNA #55
RFU
20000
15000
10000
5000
HPV18
NTC
20
40
60
Cycle
f
Primer set G | crRNA #55
RFU
20000
15000
10000
5000
HPV18
NTC
20
40
60
Cycle
g
Primer set B | crRNA #51
RFU
20000
15000
10000
5000
HPV18
NTC
20
40
60
Cycle
h
Primer set C | crRNA #53
RFU
20000
15000
10000
5000
HPV18
NTC
20
40
60
Cycle

**Fig. S1 | Screening for the best-performing set of primers/crRNAs (real-time data).** Eight primers were screened with the CRISPRD reaction for detecting HPV18. Plotted is the mean fluorescence of n=3 technical replicates and standard deviation. crRNA and gRNA are used interchangeably in the study.

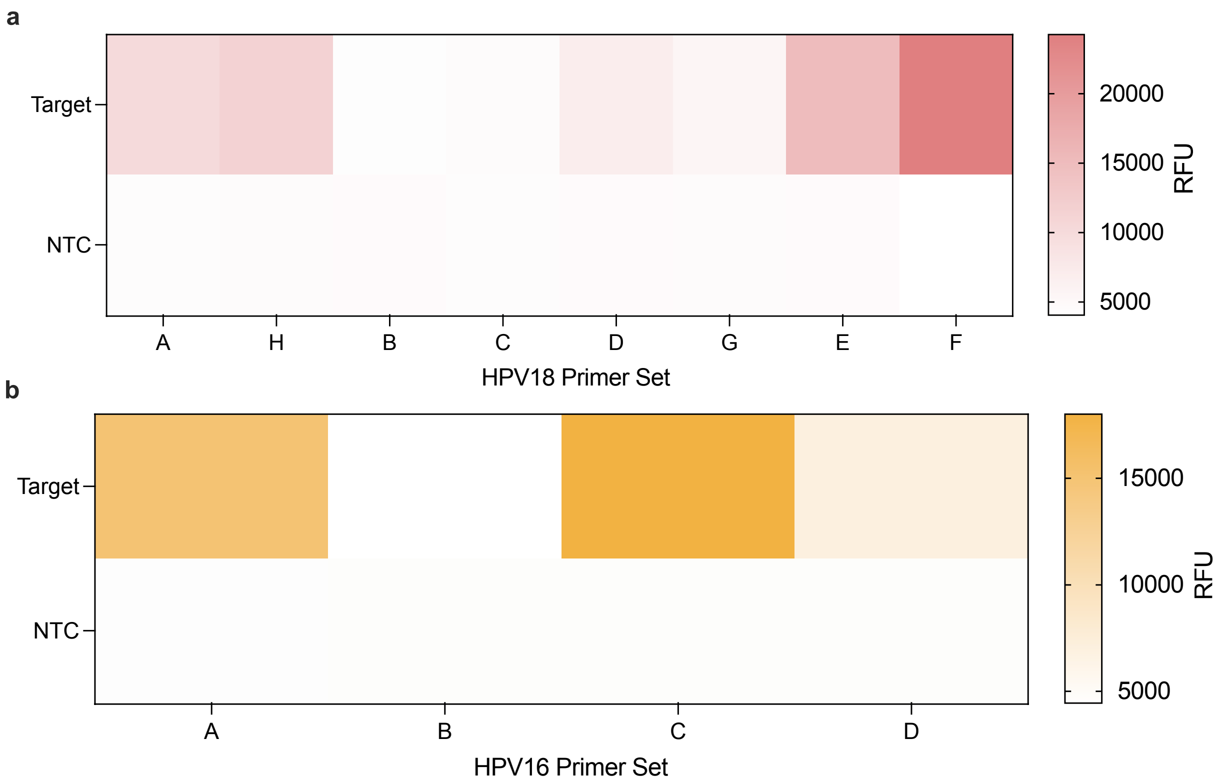


**Fig. S2 | Primer screening for HPV16 and HPV18 (Endpoint).** Endpoint fluorescence of a) 8 primer sets for HPV18 and b) 4 primer sets for HPV16 screened using the CRISPRD reaction. Plotted is the mean end point fluorescence of n=3 technical replicates and standard deviation.

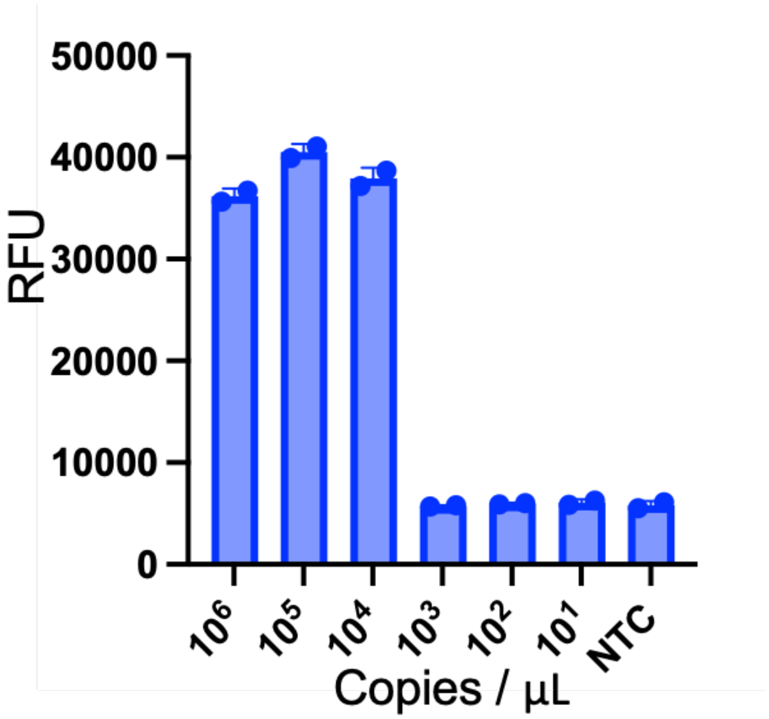


**Fig. S3 | LoD using primer set A for HPV18.** Primer set A was tested with synthetic dilutions of HPV18 DNA ranging from 0 to 1 million copies per µL. Plotted is the mean end-point fluorescence Of incubation in the CRISPRD reaction after two hours (n=3).

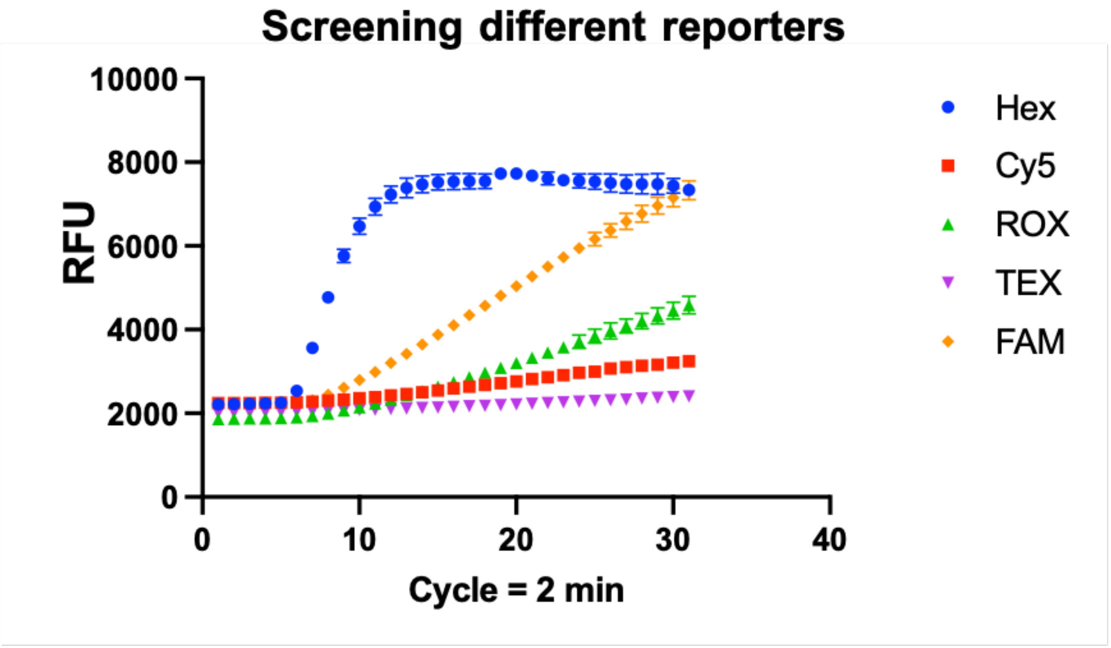


**Fig. S4 | Reporter screening.** The same reporter (Poly (rArG)) carrying different fluorophores was screened to check the sensitivity of each fluorophore.

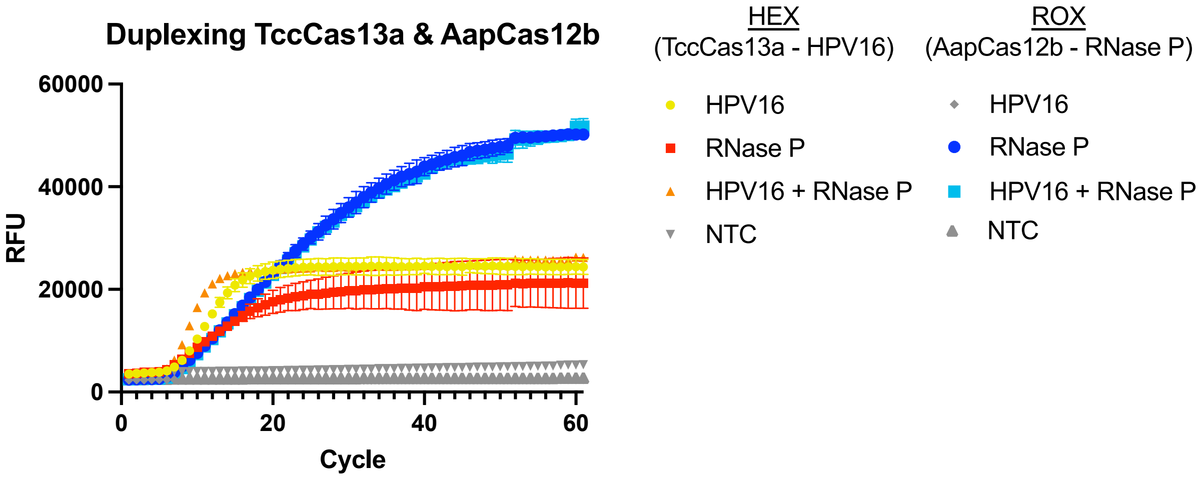


**Fig. S5 | Duplexing AapCas12b with TccCas13a.** A CRISPRD duplex reaction for detecting HPV16 and RNase P with TccCas13a and AapCas12b, respectively.

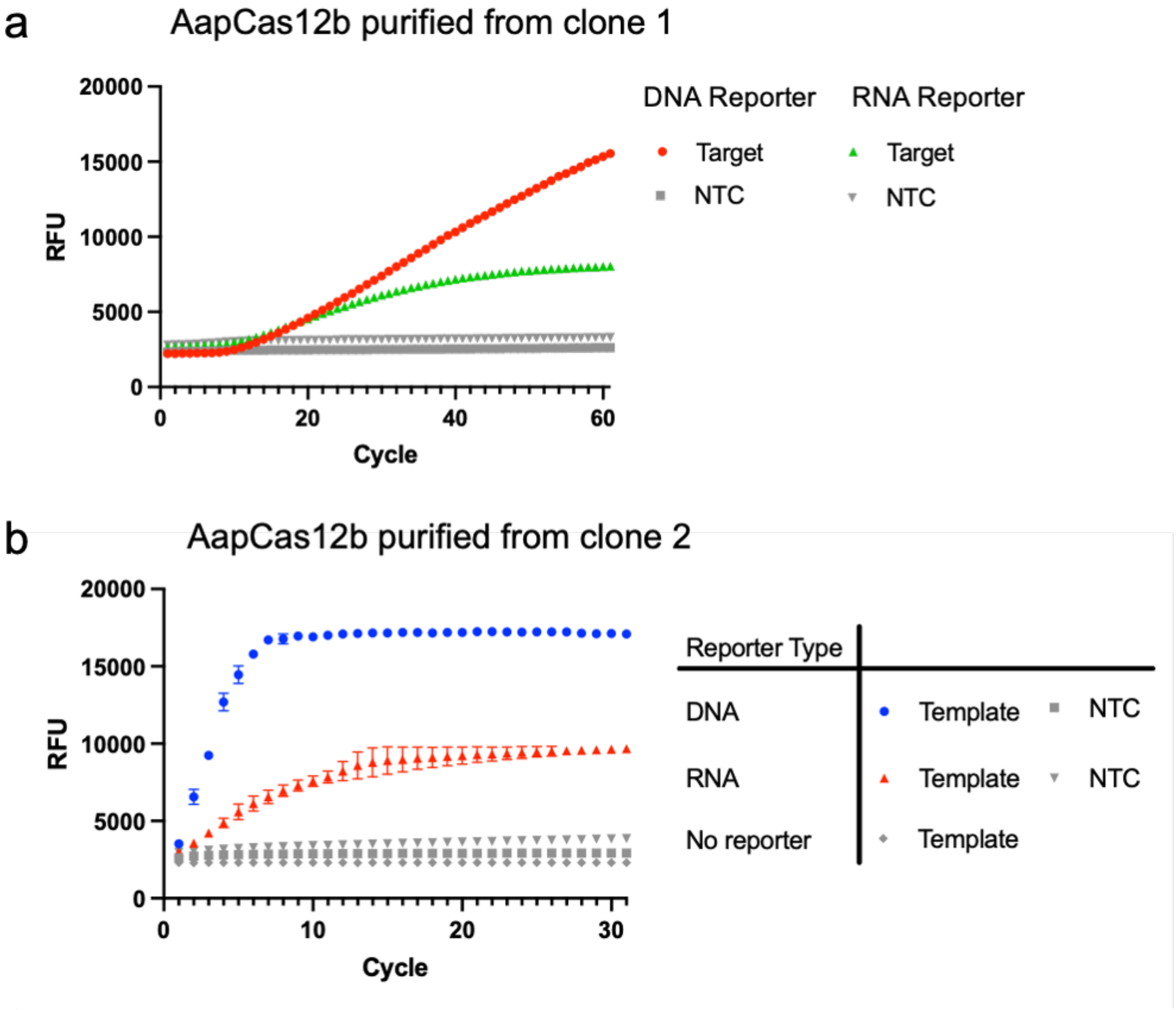

**Fig. S6 | Testing the collateral cleavage activity of AapCas12b purified from two different clones.** Two different clones of AapCas12b were tested to check their activity on an RNA reporter (poly (rArG)) compared to a DNA reporter (Poly T). Plotted is the mean real-time fluorescence data (n=3) and standard deviation.

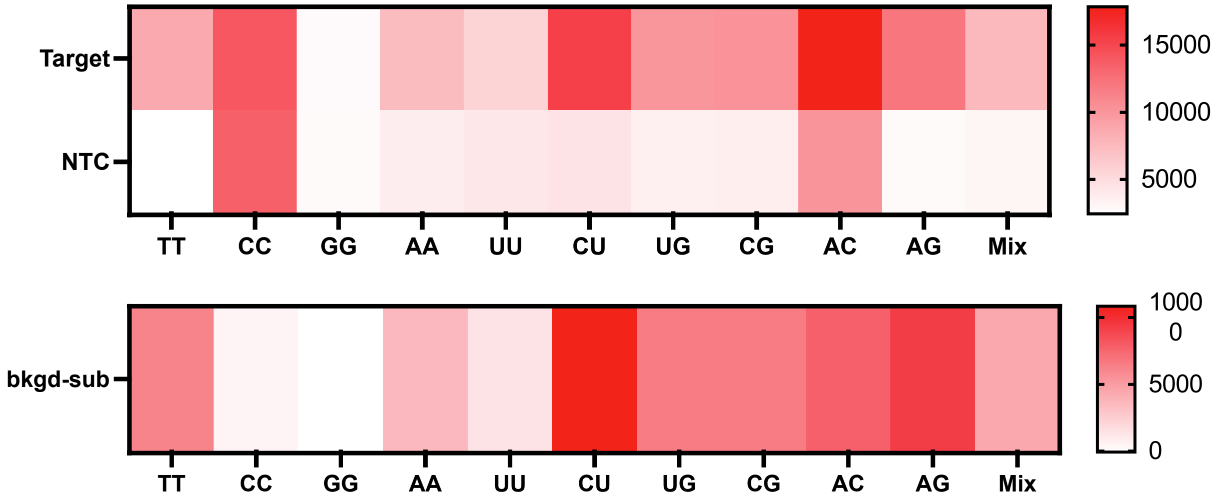


**Fig. S7 | Activity of AapCas12b on RNA reporters with different nucleotide compositions.** Activated AapCas12b RNPs were incubated with different RNA reporters. The DNA reporter is indicated as TT; the rest are RNA reporters. Plotted is the end-point fluorescence after a 1-hour incubation (n=3).

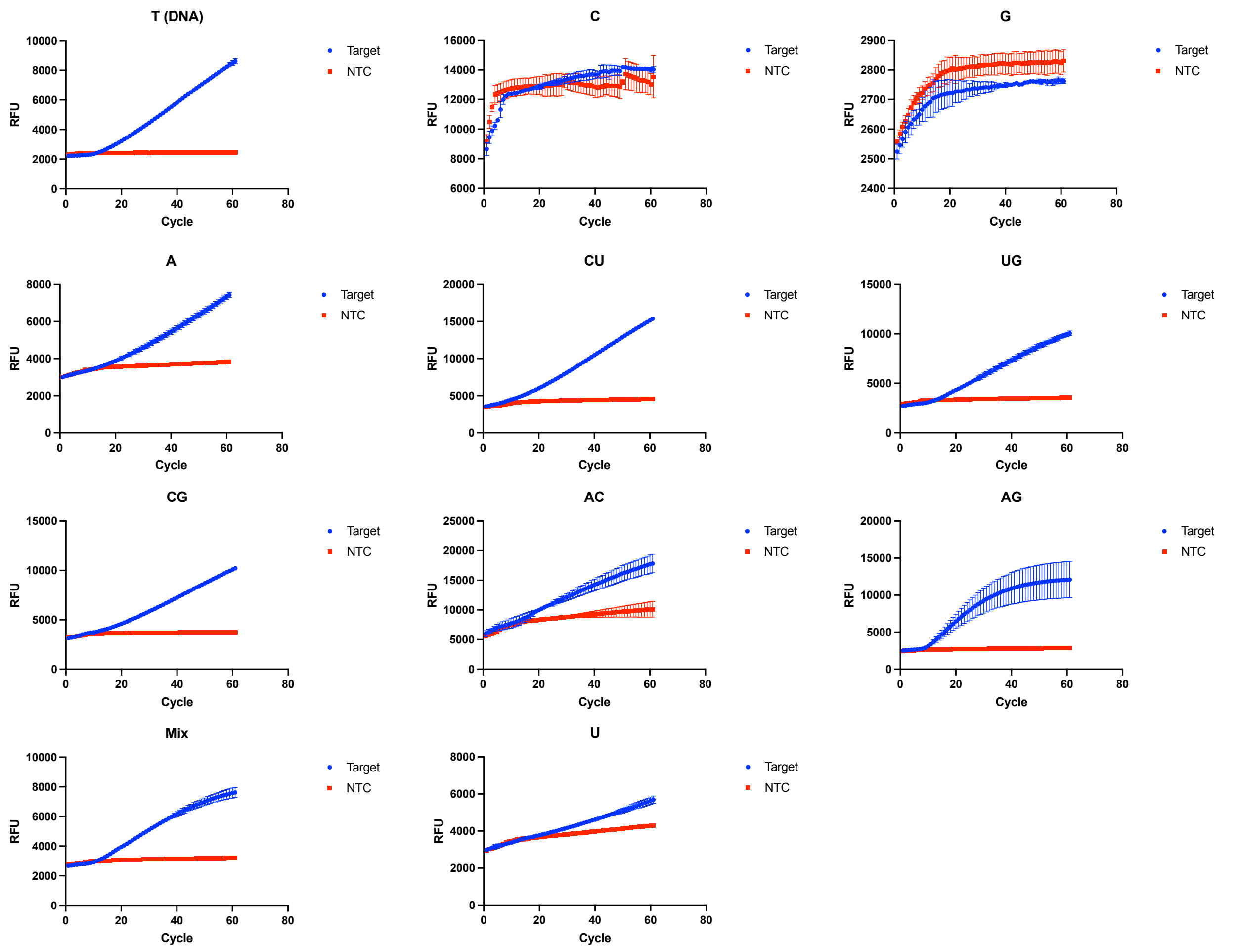


**Fig. S8 | Reporter screening with AapCas12b real-time data.** Activated AapCas12b RNPs were incubated with different RNA reporters. The DNA reporter is indicated as TT; the rest are RNA reporters. Plotted is the end-point fluorescence after a 1-hour incubation (n=3).

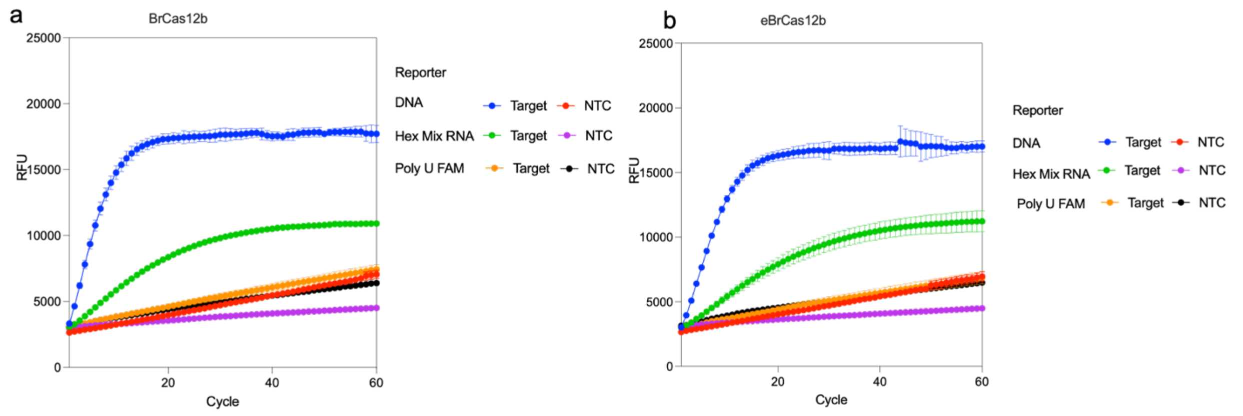


**Fig. S9 | BrCas12b cleaves RNA and DNA reporters.** Activated BrCas12b **(a)** and eBrCas12b **(b)** was incubated with RNA or DNA reporters.

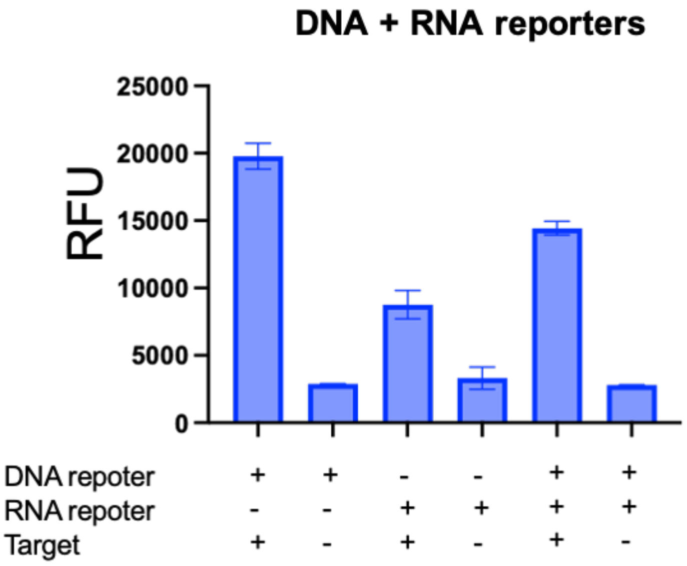


**Fig. S10 | Testing AapCas12b with RNA and DNA reporters.** Activated AapCas12b RNP was incubated with a DNA reporter, a RNA reporter, or both reporters in equimolar concentrations. Plotted is the end-point fluorescence (n=3).
